# Navigating the Molecular Maze of Alzheimer’s Disease: Developing A Human Hippocampal Atlas to Bridge Discovery and Clinical Practice

**DOI:** 10.1101/2023.12.06.23299593

**Authors:** Pan Wang, Lifang Wang, Quyuan Tao, Zhen Guo, Ting Luo, Zhi Xu, Youzhe He, Jiayi Yu, Yuyang Liu, Zihan Wu, Bin Xu, Bufan Jin, Ying Yang, Mengnan Cheng, Yujia Jiang, Chen Tian, Huiwen Zheng, Zhongqin Fan, Peiran Jiang, Yue Gao, Juanli Wu, Shengpeng Wang, Bing Sun, Yanrong Wei, Zheng Fang, Junjie Lei, Benyan Luo, Huiying Wen, Guoping Peng, Yuanchun Tang, Xinhui Su, Catherine Pan, Keqing Zhu, Yi Shen, Shiping Liu, Aimin Bao, Jianhua Yao, Jian Wang, Xun Xu, Xiaoming Li, Longqi Liu, Shumin Duan, Lei Han, Jing Zhang

## Abstract

Alzheimer’s disease (AD), the primary cause of dementia worldwide, persists as an enigma, with the specific molecular and cellular mechanisms driving cognitive decline still eluding understanding. To aid in unraveling this enigma, we generated a spatial transcriptomic atlas of the human hippocampus with single-cell resolution, leveraging integration of spatial mapping with single-cell transcriptomic profiles from six individuals with or without AD. Utilizing this complex and multi-dimensional data, the atlas pinpoints AD-associated transcriptomic changes with spatial specificity, such as a pronounced elevation of inflammatory responses and energy metabolism in fimbria and CA4, respectively. AD also caused alterations of the cellular composition in hippocampal subregions, even around the amyloid-beta (Aβ) plaques. Finally, our investigation uncovered pathways intricately linked to synthesis and secretion of extracellular vesicles (EVs). Within these altered pathways, we identified several corresponding proteins, including CCK and PMP2, that are readily detectable in human blood, holding promise for aiding AD diagnosis in clinical settings.

## Introduction

Alzheimer’s disease (AD), the most common form of dementia, is characterized by the accumulation of amyloid-beta (Aβ) plaques and neurofibrillary tangles (NFTs) in the brain, which contributes to progressive deterioration of brain function ^1–3^. However, despite being considered the hallmark of AD pathology and being the subject of decades of research, therapeutic approaches aimed at reducing these pathological proteins have demonstrated limited effectiveness and often result in severe side effects ^4,5^. Therefore, identifying novel pathological mechanisms and tissue-specific markers is crucial for AD diagnosis and treatment.

The advent of single-cell sequence and spatial genomics technologies has revolutionized our understanding of complex tissues in both healthy and diseased states. These powerful tools have begun to unravel the cellular vulnerabilities and molecular alterations associated with AD ^6,7^, but to date, their application has primarily focused on the cortex ^8–11^, missing out the hippocampus. The hippocampus, crucial for memory, navigation, and cognition, stands notably vulnerable to the early impairments associated with AD ^12–15^.To address this gap in knowledge and uncover the molecular underpinnings of AD, we focused on the research of the human hippocampus. By delving into this vulnerable region, we can seek answers to fundamental questions, including: 1) What distinguishes individuals without cognitive impairment from those who later develop dementia? 2) Which specific cell types are more susceptible to Aβ burden? 3) Which genes and pathways exhibit significant alterations in response to heightened levels of AD pathology? And 4) What genes and proteins are altered in the human hippocampus that could serve as valuable biomarkers for AD diagnosis?

In the present study, we began by reporting the insights gained from a comprehensive spatial single-cell molecular atlas of the human hippocampus. We reported transcriptomic differences associated with AD pathology, and depicted their distribution across the healthy control (HC) and AD hippocampus as an atlas of differentially expressed genes and altered cellular signatures. Further, we identified Aβ-associated alterations in microglia and astrocytes, and examine regulation of functionally linked pathways such as genes involved in synaptic functions, as well within the electron transport chain. Furthermore, to demonstrate the utility of this atlas in translation to clinical needs, we linked significantly differentially expressed genes (DEGs) and their corresponding proteins, specifically CCK and PMP2, carried by extracellular vesicles (EVs). These markers, selected based on their alterations observed in the atlas were then developed as peripheral blood biomarkers to distinguish AD from HC, thereby uncovering a link between early changes in the central nervous system (CNS) and peripheral blood of AD pathology.

By harnessing the power of single-cell technologies, our study provided a reference of intricately human hippocampus to unravel the mysteries of AD and pave the way for a future free from its devastating effects.

## Results

### Multimodal spatially-resolved transcriptomics in human hippocampus

Donors with AD neuropathologic changes (AD) and controls was characterized in a coordinated fashion to allow a joint analysis of donor demographics, clinical history (**Table 1**), neuropathology, single-nucleus RNA sequencing (snRNA-seq), and spatiotemporal enhanced resolution omics sequencing (Stereo-seq) to profile spatial and molecular features of cells within hippocampus (**Figure 1A**). Only autopsy samples with postmortem interval (PMI) < 12 hours, meeting stringent criteria of quality control (QC) procedures suitable for exceptionally high-quality analyses using snRNA-seq and Stereo-seq ^16^, were included in the investigation.

**Figure 1.**
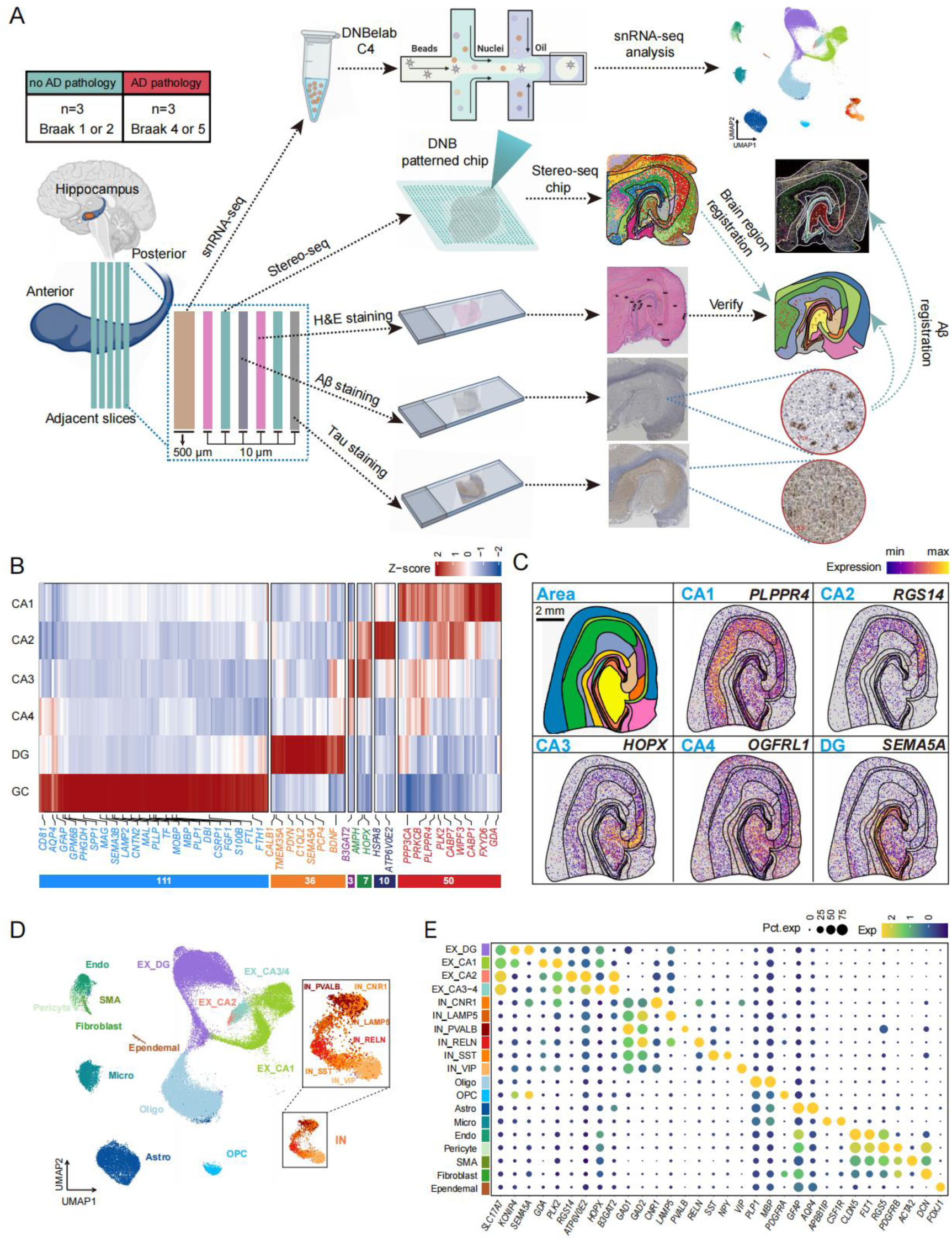
Multimodal spatially-resolved transcriptomics in human hippocampus. **(A)** Schematic diagram of experimental design. snRNA-seq, Stereo-seq, HE staining, and immunohistochemical staining were applied in this study with different adjacent slices. Subregion parcellation was conducted by integrating spatial gene expression and H&E staining for subregion specific genes screening. SnRNA-seq were used to register and annotate spatial cell map. The spatial information of Aβ was registered to Stereo-seq for analyzing gene expression and cell distribution changes surrounding Aβ. **(B)** Heatmap of subregion-specific genes in hippocampus. CA: Cornu Ammonis; DG: dentate gyrus; GC: Glial cell region. **(C)** Spatial profile of representative subregion-specific genes. **(D)** Uniform manifold approximation and projection (UMAP) visualization of hippocampal cell types. **(E)** Dot plot depicts the expression of marker genes in annotated cell clusters.

**Table 1.** Summary metrics from the CNBB cohort across AD Neuropathological Change.

| Group | AD1 | AD2 | AD3 | Control1 | Control2 | Control3 |
| --- | --- | --- | --- | --- | --- | --- |
| Sex, male: female | F | M | F | F | M | F |
| Age at Death | 84 | 73 | 93 | 65 | 76 | 94 |
| Post Mortem Interval (Minutes) | 140 | 570 | 225 | 149 | 354 | 153 |
| Brain pH | 6.55 | 8.3 | 6.81 | 6.67 | 6.74 | 6.34 |

We first collected detailed, single-cell level gene expression information using snRNA-seq. To accomplish this, 500-μm thick tissue sections of human hippocampus were dissociated and nuclei were sequenced. Each hippocampal sample yielded six adjacent 10-µm sections. Two of these sections were used for Stereo-seq (2 chips per sample), while the remaining four were used for hematoxylin and eosin (H&E) staining, to reveal the structure, and immunohistochemical staining of Aβ and phosphorylated Tau, respectively, to identify the locations of pathological deposits of proteins within the region (**Figure 1A; Figure S1A and S1B**). The immunohistochemical results displayed accumulated Aβ plaques in AD but the plaque was undetectable in control (**Figure S1A**). Simultaneously, positive signal of Tau protein was remarkably enhanced in AD compared to those in control (**Figure S1B**). These results confirmed the reliability of pathological diagnosis in this study.

In total, we collected 12 slices from 3 AD and 3 control donors for generating Stereo-seq data (**Table 1**). The quality of these data was assessed in the form of bin100 size (100 × 100 DNB spots, dimension size: 50 µm × 50 µm). On average, we achieved 2537 genes/bin100 and 6723 unique molecular identifiers (UMIs)/bin100 (**Figure S1C**), suggesting high quality of these spatial transcriptomic data. Since parcellation of mouse tissue have been achieved with Stereo-seq^17^, we performed BayesSpace clustering with bin100 dataset. The delineated subregions identified by Stereo-seq largely overlapped with that determined by H&E parcellation based on cell distribution pattern within the hippocampus (**Figure S1E**). After configuration of subregional parcellation in all slices, we obtained a series of subregion-specific genes. Our analysis revealed a total of 6 well-defined anatomical subregions within the hippocampus, including the Cornu Ammonis1 (CA1), CA2, CA3, CA4, dentate gyrus (DG) including molecular layer of DG (ML.DG), DG, and polymorphic layer of DG (PL.DG), and glial cell region (GC) including FAS (fimbria, alveus, and stratum oriens (SO)) and SLRM (stratum lucidum (SL), stratum radiatum (SR), stratum moleculare (SM)) (**Figure 1B**). Established marker genes, such as *PLPPR4* for CA1, *RGS14* for CA2, *HOPX* for CA3, *OGFRL* for CA4, *SEMA5A* for DG and *PLP1* for GC were visually depicted in all 12 parcellated slices (**Figure 1C and Figure S2**). These slices exhibited excellent reproducibility in expression of subregion-specific genes (**Figure S1H**), the implying reliability of parcellation in this work. Based on parcellation, remarkably high number of genes and UMIs were found in DG with no detectable differences in capture of genes between the two groups in each subregion (**Figure S1D**).

Aside from Stereo-seq, we also performed snRNA-seq and harvested a total of 60,249 qualified cells (1507 genes/cell and 2745 UMIs/cell) after removing cells with low quality (**Figure S1F**). We clustered and annotated all these cells into 11 main cell types (**Figure 1D**), including excitatory neuron (EX) (*SLC17A7*), inhibitory neuron (IN) (*GAD1*), oligodendrocyte (*PLP1*), oligodendrocyte progenitor cells (OPC) (*PDGFRA*), astrocytes (*GFAP*), microglia (*C1QB*), endothelial cells (*CLDN5*), pericytes (*PDGFRB*), ependymal cells (*FOXJ1*), smooth muscle cell (SMA) (*ACTA2*) and fibroblasts (*DCN*). Additionally, EXs were further categorized into 4 subtypes, including: EX_DG cells (*SEMA5A*), EX_CA1 cells (*GDA* and *PLK2*), EX_CA2 cells (*RGS14* and *ATP6V0E2*) and EX_CA3/4 cells (*HOPX* and *B3GAT2*). INs were classified into 6 subtypes, including: IN_CNR1 (*CNR1*), IN_LAMP5 (*LAMP5*), IN_PVALB (*PVALB*), IN_RELN (*RELN*), IN_SST (*SST*) and IN_VIP (*VIP*) (**Figure 1E**).

### Transcriptional differences in hippocampal subregions between AD and control

To characterize the specific molecular fingerprint of the AD hippocampus, we conducted spatial domain-based differential expression analysis. A total of 1762 DEGs, including 982 upregulated genes and 780 downregulated genes were identified after comparison between AD and control.

For the upregulated DEGs, there were 183 shared genes and 799 subregion-specific genes across the 14 hippocampal subregions (**Figure 2A and S3A**). Functional enrichment analysis showed that these 183 shared genes were mainly related to metabolic process such as protein translation, immune response and energy generation (**Figure 2A**), suggesting a global change of protein metabolism, neuroinflammation and energy metabolism in hippocampus. For the 799 subregion-specific genes, various functions were enriched in each subregion. Aβ binding pathway-related DEGs were specifically enriched in CA1, indicating this area was the main deposition site of Aβ. Notably, CA4 had the highest number of DEGs among all subregions. These DEGs derived from CA4 were highly associated with mitochondrial respiratory chain related pathways. In particular, vascular associated function pathways were enriched in SM and alveus (**Figure 2A**). Consistently, representative genes in those pathways exhibited global upregulation, such as inflammation-related gene *CD74*, and subregion-specific upregulation, such as *ATP6AP2* involved in mitochondrial respiratory chain in CA4, *HK2* correlated with HIF-1 signaling in alveus (**Figure 2C**).

**Figure 2.**
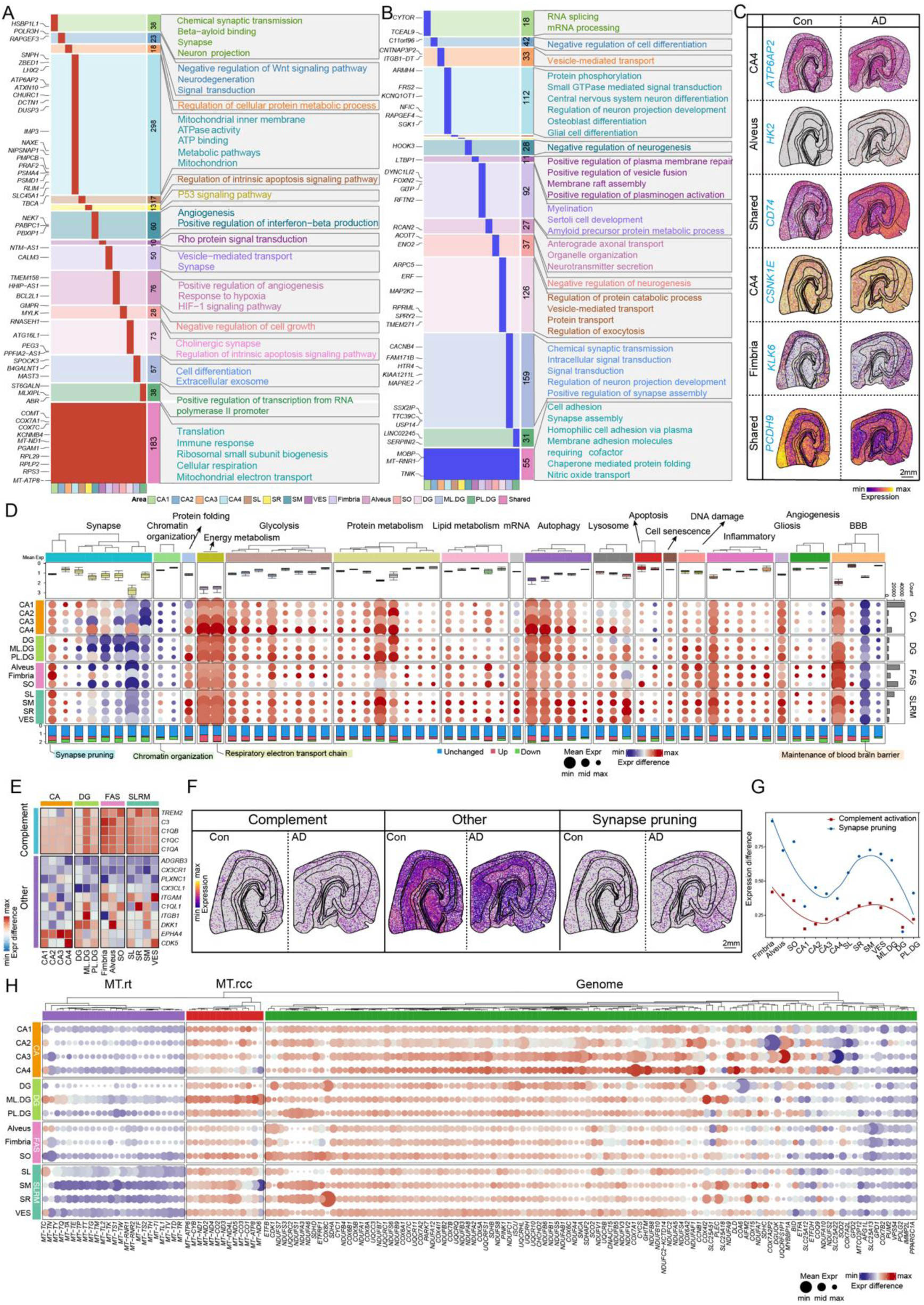
Differential gene expression and their functions based on spatial transcriptomics. **(A-B)** The results of DEG analysis and gene function annotation between AD and control in hippocampal subregions. Upregulated (red) and downregulated (blue) DEGs were shown in A and B, respectively.) The right panel of each figure showed the corresponding functions of these genes. Numbers shown in the column represented the counts of DEGs in individual subregion and the representative DEGs were shown in the leftmost box. DEGs were defined by (| log_2_FC | > 0.25 & adjusted *p*-value < 0.05). **(C)** Spatial visualization of representative shared and subregion-specific DEGs in hippocampus of AD and control. **(D)** The dot plot illustrated the differential expression of various pathways in hippocampal subregions between AD and control. The color of the top layer represented different function category. Each column represented individual pathway and the name of representative pathways were labeled on the bottom. The size of the dot indicated the average expression value of each pathway over AD and control whereas the color represented difference in expression value between the two groups. The stacked bar represented the proportion of upregulated (red), downregulated (blue) and unchanged (green) genes within individual pathway (bottom). The rightmost gray bar plot demonstrated the number of counts in bin100 data in the corresponding subregion. **(E)** Heatmap showed the expression of genes in the synaptic pruning pathway across hippocampal subregions. **(F)** Spatial visualization of the complement-related gene set score. **(G)** The curve plot showed differential expression of synaptic pruning and complement activation pathways across hippocampal subregions. **(H)** Dot plot showed the expression difference of genes related to respiratory electron transport chain across hippocampal subregions. Each column represented a gene. The meaning represented by size and color of the dots was coincide with that in Figure 2D. **(G)** The scatter plot showed gradient changes in gene expression along the CA2-CA1 axis. The y-axis indicated the average expression, and the blue line represented the linear regression of these scatters.

For the downregulated DEGs, there were 55 shared genes and 725 subregion-specific genes across the 14 hippocampal subregions (**Figure 2B and S3C**). Functional annotation showed the shared genes were mainly related to cell adhesion, protein folding and synapse assembly (**Figure 2B**). Genes related to cell adhesion, such as *PCDH9*, consistently exhibited an overall downregulation in spatial visualization (**Figure 2C**). Subregion-specific genes were mainly identified in CA4, fimbria, ML.DG, and DG. Notably, the downregulated DEGs in CA4 and DG were primarily associated with protein phosphorylation (e.g., *CSNK1E*) and protein catabolism. Myelination and amyloid precursor protein metabolic process associated functions were enriched in fimbria. A gene, *KLK6*, from these two pathways was shown to be downregulated in oligodendrocyte enriched subregions including Alveus and fimbria (**Figure 2B and Figure 2C**).

Beside elevation of inflammation in context of AD as shown above, AD progression also accompanied with alteration in cell senescence, autophagy, metabolism, angiogenesis, apoptosis, lysosome, protein folding, gliosis, chromatin organization and synapses ^9^. We, thus, systematically explored AD-induced changes in those molecular pathways across hippocampal subregions. Apart from the downregulation of synaptic pathways, chromatin organization, and maintenance of blood brain barrier, most pathways were upregulated in AD. Cell senescence and apoptosis were both represented higher upregulation in SO, SM and SR. Angiogenesis was enriched in glia region and most pronounced in alveus and fimbria. In addition, dramatic upregulation of autophagy, metabolism, lysosome and protein folding was shown in CA4, indicating this subregion may be regarded as a pathological landmark of hippocampus with AD (**Figure 2D**).

Specificially, despite the majority of synaptic pathways were downregulated and this downregulation was prominent in DG (DG, ML.DG, PL.DG) and FAS (Fimbria, Alveus, SO), synaptic pruning stood out by showing an upregulation trend. It was noteworthy that the upregulation of the synaptic pruning appeared to be more prominent in the glial region, such as ML.DG, alveus, fimbria and SL (**Figure 2D**). We observed an overall upregulation of genes related to the complement system in the synaptic pruning pathway, especially pronounced in the fimbria, alveus and ML.DG region (**Figure 2E**). Spatial visualization of the complement-related gene set score further corroborates this observation (**Figure 2F**). In line with synaptic pruning, a global elevation of complement activation was detected, but showed differences in extent across subregions (**Figure 2G**).

We observed an upregulation of the energy metabolism pathways across all subregions, indicating dysfunction of energy metabolism in AD hippocampus **(Figure 2D**). The operation of energy metabolism required coordinated activities of respiratory electron transport chain gene set from both mitochondrial and nuclear genome ^9,18^. Intriguingly, a part of MT genes coding for mitochondrial respiratory chain complex (MT.rcc) were globally upregulated and especially pronounced in ML.DG, while the left mitochondrial genes that encode rRNA and tRNA (MT.rt) were downregulated (**Figure 2H**). Similarly, nuclear genes belonged to respiratory chain pathway also can be divided into two parts with opposite expression patterns in global. It was of note that the upregulation of genes from nuclear genome was more pronounced in the CA4 area (**Figure 2H**).

In addition, to unravel average changes of gene expression occurring in each anatomical subdivision, we also investigated internal spatial changes within the subdivision leveraging Stereo-seq. Two axes in SLRM and CA1 were defined respectively (**Figure S3C**). Along the CA1-DG axis, parallel with extending dendrites from the EX_CA1, in SLRM, we established a linear model based on the expression levels of each gene at each spatial layer. Through this linear model, we first identified genes exhibiting gradient expression pattern along the axis and then screened genes with distinct spatial gradient pattern in AD (**Figure S3D**), such as *BIN1, LDHA, LDHB* and *ENO1* (**Figure S3D and S3E**). Since previous studies implicated BIN1 as a risk gene for AD ^18^, we showed an opposite expression trend between AD and control, further supporting its high risk in AD. Interestingly, in AD, *LDHA*, *MT-ATP6* and *ENO1,* associated with energy metabolism ^19–21^, demonstrated opposite expression patterns relative to the control group (**Figure S3E**), implying dysfunction of energy metabolism along the axis. Along the CA2-CA1 axis, parallel to neuronal information flow in CA, we also gained genes with distinct spatial gradient pattern in AD, such as *CAMK1, CALM3, CAMK2N1* and *SLC6A7* (Figure **S3F and S3G**). As all these four genes were essential for synaptic transmission ^22–25^, pathological changes of these genes along the axis indicated abnormal neural information relay in CA1.

Collectively, these results exhibited a spatial map of AD-triggered gene expression changes in hippocampus.

### Spatial distribution changes of cell types between AD and control

To identify the spatial location of hippocampal cell types and their gene expression signature, we used an AI-assisted automatic segmentation algorithm for delineating single-cell boundary based on ssDNA staining **(Figure S4A)** ^17,26^. On average, we obtained 58,424 segmented cells/section with 572 UMIs/cell and 293 genes/cell after quality control **(Figure S4B, C)**. To annotate the segmented cells in Stereo-seq data, we applied Spatial-ID algorithm by integrating snRNA-seq with the Stereo-seq data **(Figure 3A)** ^26,27^. Using this method, we identified 19 cell types with a total number of 701,092 single cells in the hippocampus. We confirmed the reliability of cell identity by classical cell markers, correlation analysis, neighborhood enrichment analysis, and consistency of cell composition between Sn-seq and Stereo-seq, respectively **(Figure S4D-G)**. In addition, there was no obvious difference in number of UMIs/cell and genes/cell between each cell type of AD and control (**Figure S4H**). The spatial distribution pattern displayed that excitatory neuron, such as EX_CA1, EX_CA2, EX_CA3/4, and EX_DG, tended to enrich in corresponding subregions, whereas inhibitory neurons, and non-neuronal cell types were dispersed throughout the entire hippocampus, except for oligodendrocyte which mainly located in glial subregion (SLRM and FAS) (**Figure 3A-C; Figure S4J**).

**Figure 3.**
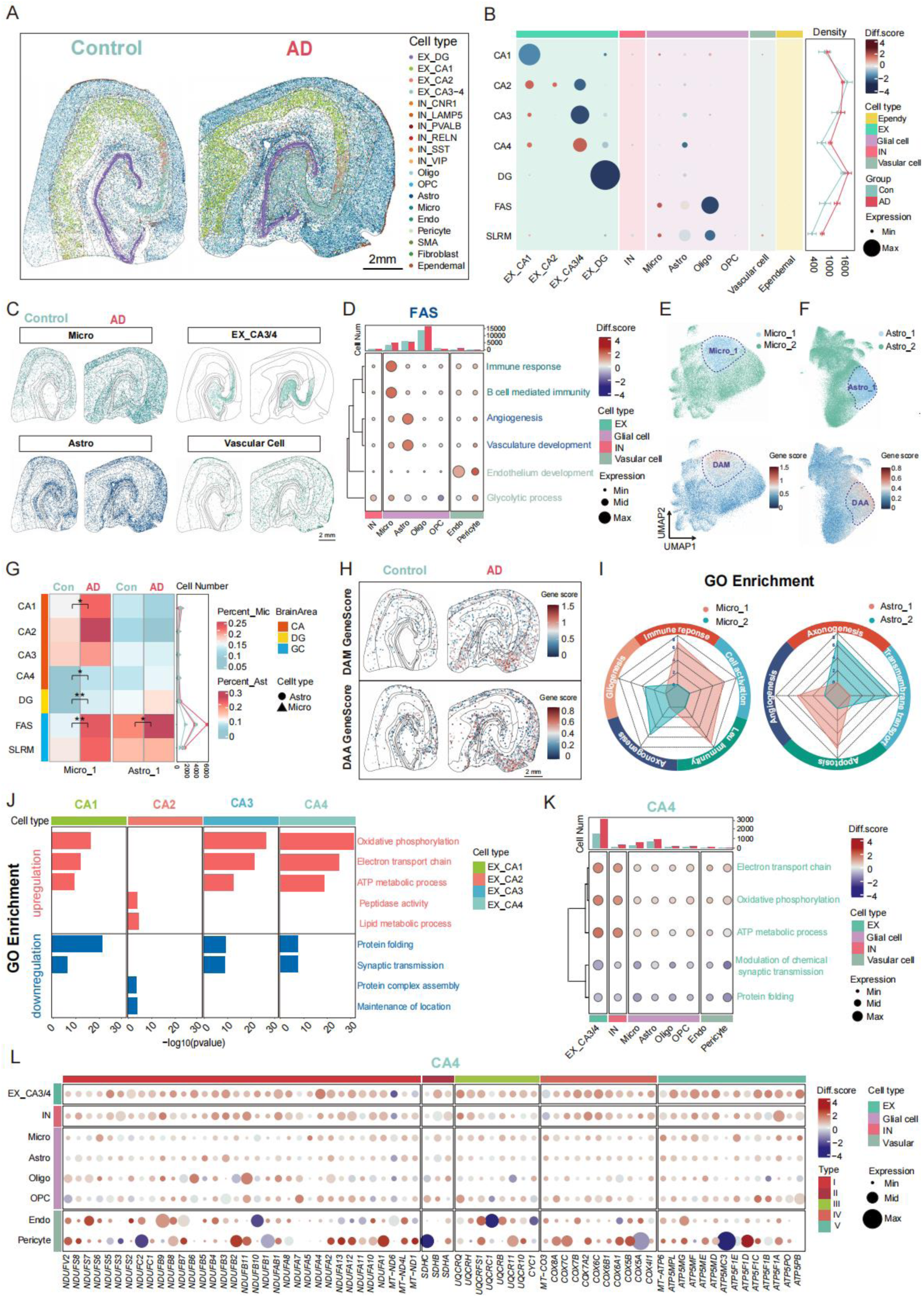
Spatial distribution changes of cell types between AD and control. **(A)** Spatial distribution of cell types in AD and control hippocampus. Scale bar: 2 mm. **(B)** Dot plot showing the proportion of each cell type change in each subregion. The spot size represented the average proportion of each cell type in each subregion over AD and control. The spot color represented increase (red) and decrease (blue) of cell proportion in AD. Diff. score, difference score. **(C)** Spatial distributions of microglia, astrocyte, vascular cell and EX_ CA3/4 in AD and control. Scale bar: 2 mm. **(D)** Dot plot showing expression change of selected enrichment pathways in all cell types of FAS subregion. Dot size represented the average expression value of each pathway genes of each cell type over AD and control whereas dot color indicated upregulation (red) or downregulation (blue) of difference value in AD. Pathways in green color, blue and light green represented top GO pathways of microglial, astrocyte and vascular specific DEG genes in AD, respectively. **(E)** UMAP visualization of microglial subclusters (top) and expression of DAM related genes in microglia subclusters (bottom). **(F)** UMAP visualization of astrocyte subclusters (top) and expression of DAM related genes in astrocyte subclusters (bottom). **(G)** Heatmap showing the proportion of DAM (micro_1) in total microglia (left) and DAA (astro_1) in total astrocyte (right) in all hippocampus subregions of AD and control. Difference between AD and control was examined by *t-test* with holm’s correction, * *p* < 0.05, ** *p* < 0.01. **(H)** Spatial distribution of DAM (Top) and DAA (bottom) related genes expression in AD (right) and control (left) hippocampus. Scale bar: 2 mm. **(I)** Radar plot showing GO functional enrichment for microglia (left) and astrocyte (right) subclusters. **(J)** Bar plot showing GO terms (rows) enriched by genes significantly upregulated (red) or downregulated (blue) in EX_CAs of AD. **(K)** Dot plot showing expression change of energy generation, synaptic transmission and protein folding pathways in all cell types of CA4. Dot size represented the average expression value of each pathway genes of each cell type over AD and control. Dot color represented upregulation (red) or downregulation (blue) of difference value in AD. **(L)** Dot plot showing the expression changes of mitochondrial complex I-V encoded genes in all cell types in CA4. Dot size represented the average expression value of each gene of each cell type over AD and control. Dot color represented upregulation (red) or downregulation (blue) of difference value in AD.

We next sought to identify AD-related changes in cell distribution within hippocampus. The cell density was varied among hippocampal subregions, ranking higher in CA2 and DG, but lower in CA1 and SLRM. This spatial pattern of cell density was consistent between AD and control, however, the proportion of each cell type in subregions was substantially changed in AD (**Figure 3B**). In line with previous report ^28–30^, we also found that EX_CA1 and EX_DG decreased in CA1 and DG of AD, respectively. Interestingly, the proportion of EX_CA2 increased in AD CA2, and that of EX_CA3/4 increased in CA4 but decreased in CA3 of AD. These results suggested that variance in proportion change of excitatory neurons might reflect their different vulnerability to AD. Since an elevation in EX_CA2 proportion in AD, this cell type might be of AD resistance ^31^, supported by enrichment of Wnt signaling pathway and lipid metabolism in EX_CA2 (**Figure S4I**). In agreement with snRNA-seq data, we also found a rise in the proportion of microglia with AD (**Figure S4G**), which is consistent with previous reports ^32,33^. Notably, in AD hippocampus, there was a pronounced increase of proportion of microglia in FAS, SLRM and CA1, whereas a slight increase of proportion of astrocyte in FAS and CA1(**Figure 3B**). In line with microglia, AD associated increase of vascular cell was shown in FAS, SLRM and CA1. Nevertheless, oligodendrocyte was dramatically reduced in FAS and SLRM (**Figure 3A-C, Figure S4J**). Since dramatic changes of cell composition in FAS, we next compared transcriptomic profile in each cell type between AD and control. Immune-related activity was pronouncedly enhanced in microglia, and vascular related pathways were remarkably upregulated in astrocyte, endothelial cell and pericyte (**Figure 3G)**. These results suggested a correlation between changes in cell ratio and their functions. To investigate whether cell state was associated with AD, we re-clustered microglia into two main subclusters, and one of them enriched in expression of disease activated microglial (DAM) genes ^32,33^(**Figure 3E**), suggesting this subcluster (micro_1) was closely related to DAM. As a result, micro_1 was designated as DAM. After calculating the ratio of DAM in total number of microglia in each subregion, we found a global increase of DAM in AD, significantly high in FAS, DG, CA4 and CA1 (**Figure 3G**). DAM gene expression in microglia also revealed enrichment of DAM in FAS (**Figure 3H**). GO enrichment analysis of DEGs between DAM and micro_2 showed that DAM was more active in immune-related response while micro_2 was associated with axonogenesis and gliogenesis (**Figure 3I**).

Meanwhile, we performed similar analysis in astrocyte, and obtained two main subclusters (astro_1 and astro_2). A gene set highly associated with disease activated astrocyte (DAA) ^32^ was specifically enriched in astro_1, so it was designated as DAA (**Figure 3F**). Using the same calculation method in microglia, we found DAA was increased in FAS (**Figure 3G**). DAA gene set expression in astrocyte confirmed the enrichment of DAA in FAS (**Figure 3H**). GO enrichment analysis of DEGs between DAA and astro_2 showed that DAA was enriched in angiogenesis and apoptosis while astro_2 was associated with axonogenesis and transmembrane transport (**Figure 3I**).

To investigate the AD-associated genes of EX_CA neurons, we conducted DEG analysis between AD and control within the same region for each EX_CA neuron cell type. Functional enrichment analysis showed that, except for EX_CA2, regardless of EX_CA cell types and, the genes upregulated in AD were all enriched for functions in ATP metabolic-related pathways, such as oxidative phosphorylation, ATP metabolic process, and electron transport chain, while genes involved in protein folding and synaptic transmission were decreased in AD (**Figure 3J**). Consistent with our comparative analysis in energy metabolism pathway, elevation of this pathway was prominent in EX_CA3/4 of CA4 (**Figure 2D, H and Figure 3J**). We wondered if inhibitory neuron and non-neuronal cell were affected in those upregulated and downregulated pathways. Intriguingly, all cell types within CA4 exhibited a coincident upregulation of energy metabolism and downregulation of synaptic transmission and protein folding, but difference in extent (**Figure 3K**). Since mitochondrial complexes were responsible for energy generation and were key components in energy metabolism, we next investigated the expression change of genes encoded mitochondrial complexes in AD. In hippocampus, expression level of mitochondrial complex I-V genes in EX_CA3/4 was remarkably upregulated. Intriguingly, EX_CA2 type cells were quite different from other cell types, showing a downregulation of mitochondrial complex I-V genes (**Figure S5**). When looking into CA4, upregulation of mitochondrial complex I-V genes remain prominent in CA3/4 and inhibitory neurons, in agreement with energy metabolism pathway (**Figure 3L**). In all, we provided a cell distribution map and simultaneously illustrated AD-triggered pathological change in cell composition within hippocampus.

### Spatial registration of Aβ plaque-associated pathological changes in AD hippocampus

To quantify Aβ load in subregions of hippocampus, we aligned hippocampal slice employed for Aβ immunohistochemical staining to the adjacent slice with parcellation. After that, Aβ plaques were successfully registered to the spatial transcriptome image (**Figure 4A and 4B**). The distribution of Aβ plaques was displayed in different subregions. Quantified analysis demonstrated that the Aβ exhibits relative high-level deposition in the CA1, CA4 and SLRM (SL, SM, SR) region despite variance of Aβ counts in individual region across three donors (**Figure 4C**), indicating heterogeneity and complexity of AD. To better characterize the molecular response to plaque deposition, we categorized three group for differential expression analysis, including Aβ enriched area, non-Aβ area (the same hippocampus subregion with high Aβ deposition) in AD slices and control area (location corresponding to Aβ enriched area in control hippocampus) (**Figure S4A**). After comparative analysis, we identified 407 DEGs around Aβ plaques. These Aβ plaque-related genes were composed of 376 upregulated genes and 31 downregulated genes (**Figure S4B**). In Aβ-enriched neuronal regions such as CA1 and CA4, the upregulated genes were primarily associated with pathways related to oxidative phosphorylation (e.g., *COX5B* and *NDUFA* family), cell adhesion (e.g., *CCN3*, *SYT11* and *TNR*), autophagy (e.g., *MAP1LC3B*, *RAB8B* and *UBB*) and regulation of neuron apoptotic process (e.g., *CLU*, *HINT1* and *BEX* family) (**Figure 4D and 4E**). This suggested that in neuronal areas, the enrichment of Aβ to some extent may lead to neuronal damage and loss. Besides, in Aβ-enriched glial cell regions like SLRM, the upregulated genes are predominantly involved in processes like inflammation, glial cell proliferation and protein folding (**Figure 4F**). This may imply the presence of significant neuroinflammatory responses in SLRM, and the upregulation of these genes likely aimed to response to inflammation and promote immune reactions. These results further hint the possibility that the Aβ deposition could affect local neural circuit around plaques.

**Figure 4.**
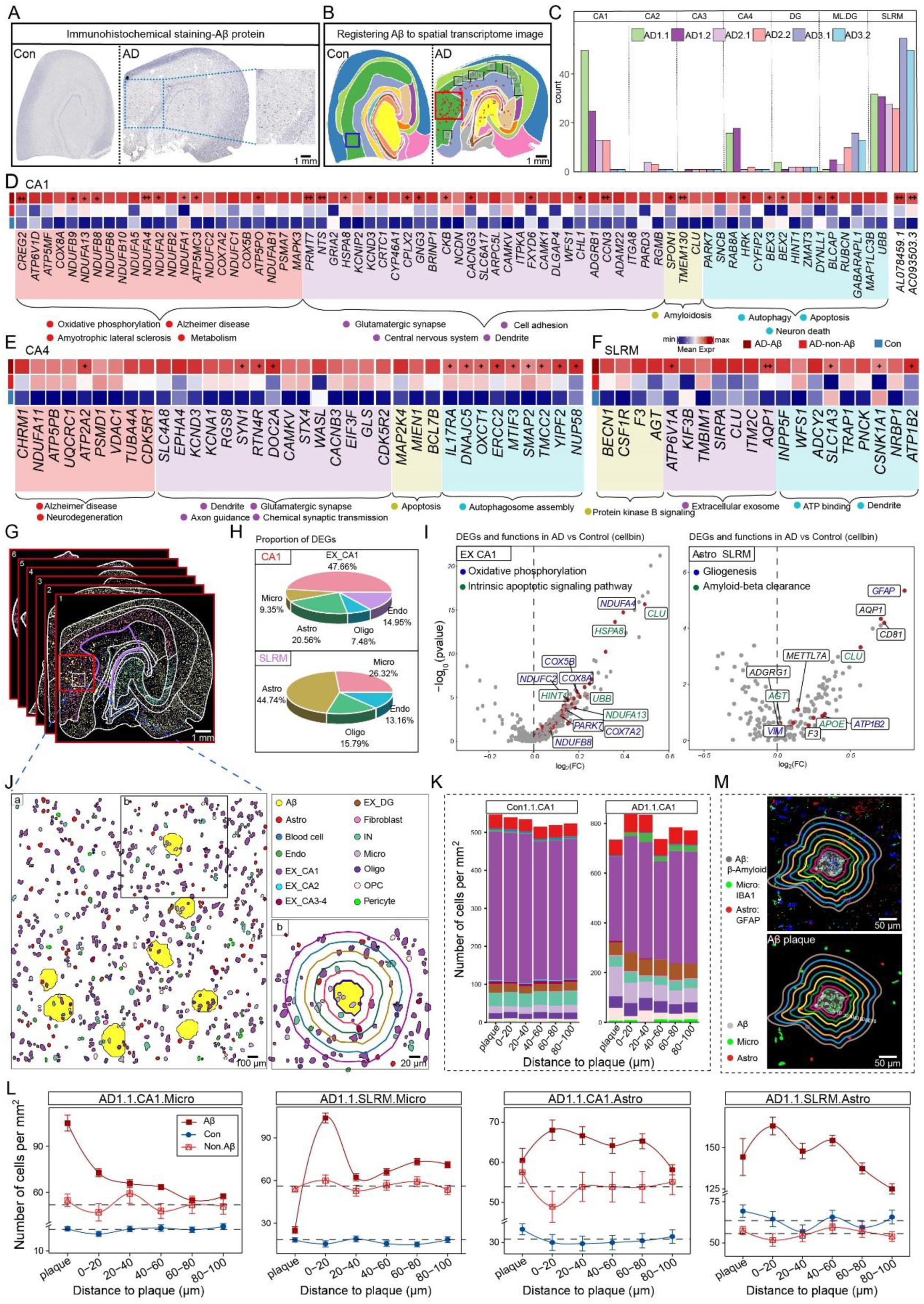
Aβ plaque-associated gene and cell type density change. **(A)** Images represented the immunostaining of Aβ protein in control (left) and AD (right) samples. **(B)** Registrating position of Aβ protein to spatial transcriptomics images. The images showed the Aβ plaque (red dot) distribution in control (left) and AD (right). In the AD group, the red box represented Aβ-dense distribution areas (AD-Aβ squares), and the gray boxes represented non-Aβ-dense distribution areas (AD-non-Aβ squares). In the control group, the blue box represented the region corresponding to the Aβ-dense distribution areas in AD. **(C)** Bar plot showing statistics of Aβ counts per anatomical region on the six AD chips. **(D-F)** Heatmap showing representative GO terms enriched among Aβ plaque-related genes across the (D) CA1 and (E) CA4 and (F) SLRM. Aβ plaque-related genes were identified through the DEG analysis (see methods). +: log_2_FC > 0.15; ++: log_2_FC > 0.25. **(G)** Representative spatial cell map embedding with Aβ plaques in AD hippocampus. Boundary lines of Aβ enriched area in CA1 and SLRM were colored in red and purple, respectively. Scale bar: 1 mm. **(H)** Pie plot showing proportions of DEGs in different cell types among the Aβ plaque-related genes within CA1 and SLRM regions. **(I)** Volcano plots showing the DEGs and their functions in EX_CA1 (within CA1) and Astro (within SLRM) in AD compared with control group. The gray dots represented DEGs of the cell type and the red dots represented the intersection of the cell type DEGs and Aβ plaque-related genes. **(J)** Schematic diagram illustrated representative spatial distribution of cell types compositions around Aβ plaque in CA1 (Zoomed in from white box in Figure 4G). The spatial patterns analysis of cell type compositions around the Aβ plaque in box b. Scale bar: (a) 100 μm; (b) 20 μm. **(K)** Stacked bar plot showing the density (number of cells per mm^2^) of each major cell type at different distance intervals (0-20, 20-40, 40-60, 60-80 and 80-100 μm) to the Aβ plaque. The density of each major cell type in control CA1 was included as the reference for comparison. The color for each cell type was coincident with Figure 4J. **(L)** Line plot showing cell density change around Aβ plaque in different distance intervals. Solid red boxes represented AD-Aβ concentric circles. Red hollow boxes represented the AD-non-Aβ concentric circles. Blue circles represented position concentric circles in control CA1 and SLRM. **(M)** Immunofluorescent staining of Aβ plaques (β-Amyloid, gray), microglia (Iba-1, green), astrocyte (GFAP, red), and nuclei (DAPI, blue) in Aβ-absent and Aβ-enriched area. Concentric rings at 10 μm intervals displayed the distribution of different cell types around Aβ at varying distances. Scale bar: 50 μm.

To explore whether Aβ deposition affect gene expression in cell types around the plaque, we compared cell types in Aβ-enriched subregions (CA1, CA4 and SLRM) with ones in corresponding subregions from control hippocampus. We obtained DEGs derived from the comparison of individual cell types and performed intersection between these DEGs and the Aβ plaques-related genes characterized by the above bin100 analysis. In the CA1 and CA4 region, Aβ plaques-related genes were primarily derived from excitatory neurons (CA1, 47.66%; CA4, 40%), and these genes were mainly involved in the regulation of oxidative phosphorylation and intrinsic apoptosis signaling pathways (**Figure 4H, I and S4C**). In the SLRM region, Aβ plaques-related genes mainly originated from astrocytes (44.74%), and these genes played essential roles in gliogenesis and Aβ clearance functions (**Figure 4H and I**).

Using ssDNA and mRNA images, we generated a cellular-resolution spatial cell atlas of the hippocampus in conjunction with precise localization of AD-related histopathology (**Figure 4G and J**). Furthermore, we assessed the effect of Aβ plaques on the composition of their surrounded cell types. To quantify cell type distribution in the spatial relationship to Aβ plaque, we set five concentric circles spaced 20 μm apart with the plaques as the center. We systematically calculated the density of different cell types in each region (**Figure 4J**; methods).

Among 13 major cell types within CA1, microglia and vascular cells in AD displayed relative enrichment around the plaques compared to their cell density with control, whereas the density of inhibitory neuron reduced, which can be repeated in replicates. Interestingly, microglia remained arising in density, however, astrocyte went down in CA4 of AD (**Figure 4K and S4E**). In SLRM, a non-neuronal cell enriched subregion, microglia and vascular cell increased accompanied with a slight reduction in astrocyte. Moreover, in AD, the CA1 region showed that aside from excitatory neurons, microglia and astrocytes were the predominant cell types around the plaques. We analyzed the changes in the density of these cell types at different distances from Aβ plaques in the CA1 and SLRM brain regions. In control, the density of microglia around the plaques at various distances remained relatively stable. However, in AD, the density of microglia around the plaques showed a declining trend with a distance-dependent manner (**Figure 4K and 4M**). The clustering of microglial around the plaques, as revealed by spatial transcriptomics, is consistent with previous research ^34^. In particular, microglia tended to be accumulated within 40 μm and dramatically reduced outside 40 μm, whereas astrocyte rose to a plateau quickly and declined beyond 60 μm **(Figure 4M**). We next validated the distribution of microglia and astrocyte around Aβ in the hippocampus of AD through immunofluorescent staining, which showed coincident spatial distribution pattern alternation of microglia or astrocyte with Stereo-seq data (**Figure 4L**).

Here, we first revealed the correlation of Aβ plaque and spatially pathological changes of gene expression and cell composition in human hippocampus.

### Detecting the DEG proteins in the blood EVs for precise diagnosis of AD

One of the primary applications of defining AD-associated alterations with single-cell resolution in spatial transcriptomics is the identification of novel region- and cell-specific biomarkers, with direct implications for clinical utility. Given the diverse changes in the functions of the hippocampus, our attention pivoted toward the processes of the synthesis and transport of EVs, with the objective of detection of these EVs in human peripheral to facilitate their potential for clinical translational applications. To be noted that cellular component (CC) of GO enrichment analysis of cell specific DEGs between AD and control in snRNA-seq data showed that DEGs in AD were enriched in vesicle-associated pathways, such as transport vesicles and coated vesicles (**Figure 5A and 5B**), suggested changes in the synthesis and secretion of EVs in hippocampus with AD.

**Figure 5.**
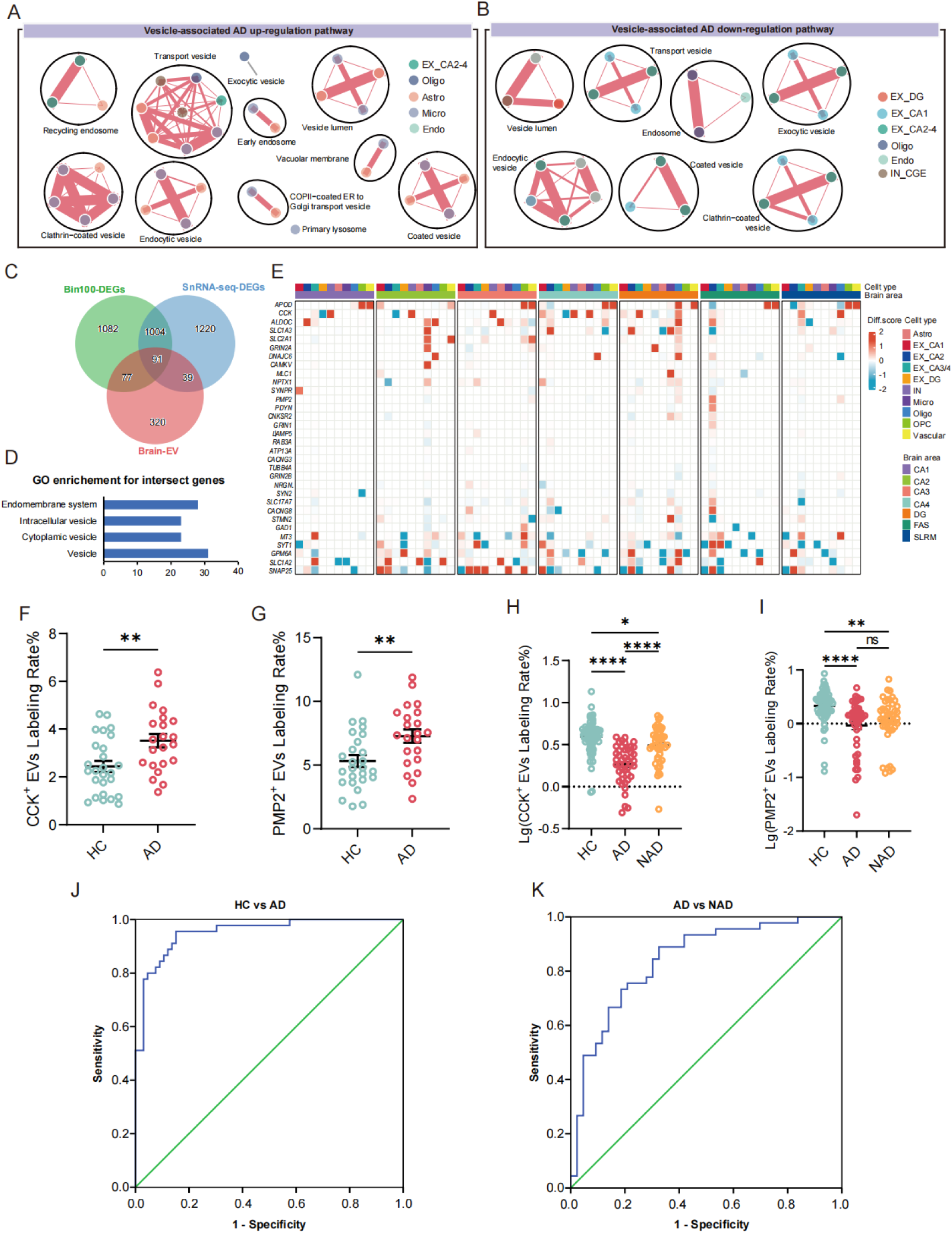
Detecting the DEGs relative proteins carried by EVs from discovery to clinical Translation. **(A-B)** Gene enrichment analysis showing upregulated (A) and downregulated (B) vesicle associated pathways in each cell type with snRNA-seq data. **(C)** The Venn diagram showing intersection of brain-derived EV genes (Brain-EV), DEGs from bin100 of Stereo-seq (Bin100-DEG) and snRNA-seq (snRNA-seq DEG). **(D)** Bar plot showing the enriched GO terms of the intersect genes in (C). **(E)** The heatmap showing different expression value (AD minus control) for enriched pathway genes in each cell type of each subregion. **(F)** The percentage of CSF CCK^+^ EVs labeling rate. **(G)** The percentage of CSF PMP2^+^ EVs labeling rate. **(H)** The percentage of plasma CCK^+^ EVs labeling rate. **(I)** The percentage of plasma PMP2^+^ EVs labeling rate. **(J)** Integrative model including CCK^+^ and PMP2^+^ EV markers distinguished AD from HC. **(K)** Integrative model including CCK^+^ and PMP2^+^ EV markers distinguished AD from NAD. * *p* < 0.05 compared to HC. ** *p* < 0.01 compared to HC. *** *p* < 0.001 compared to HC. **** *P* < 0.0001 compared to HC. Kruskal-Wallis test followed by Dunnett’s multiple comparisons test.

To figure out whether EVs, originated from brain, can be detected in peripheral blood for distinguishing AD from control, we first collected DEGs from head to head comparison in each subregion with bin100 Stereo-seq data and in each cell type with snRNA-seq data (**Figure S7A and S7B**). After that, we intersected above two DEG lists with brain-derived EVs in exoRBase (http://www.exorbase.org/exoRBaseV2/search/toIndex) and finally got 91 AD associated and brain-derived EVs DEGs (**Figure 5C**).

GO enrichment analysis showed the 91 EVs-DEGs were associated with the endomembrane, intercellular vesicle, cytoplasmic vesicle and neuron projection (**Figure 5D**). The expression alterations of some representative EVs-DEGs in each cell type and hippocampal subregions were showed in the heatmap, where CCK stood out in IN in CA (**Figure 5E**). In order to screen for peripheral blood biomarkers which can be conveniently used for clinical precise diagnosis of AD, we selectively labeled a range of proteins coded by these EVs-DEGs on plasma EVs, among which CCK and PMP2 performed well in discriminating AD from HC in a small cohort (**Figure 5SC-5SD**). Meanwhile, we confirmed that CCK and PMP2 can be detected on EVs from the cerebrospinal fluid (CSF), and both of them indeed can effectively distinguish between AD and HC (CCK^+^ EVs, HC VS AD, \*\**p*<0.01; PMP2^+^ EVs, HC VS AD, \*\**p*<0.01) (**Figure 5F and 5G**). The EVs size and distribution (**Figure 5SE**), morphology (**Figure 5SF**), and the assay specificities and dilution linearity (**Figure 5SG-5SK**) were also confirmed. A single reference plasma sample was run in duplicate, and repeated over 5 days, to demonstrate the day-to-day stability. Coefficients of variation for intra- and inter-day assay comparisons were ≤ 10% for all markers. Next, we conducted a cohort study to evaluate the performance of CCK and PMP2 for distinguishing AD from HC. Notably, the ratio of CCK^+^ and PMP2^+^ EVs were lower in AD and NAD compared to HC (**Figure 5H and 5I**). It is worth noting that the ratio of both CCK^+^ and PMP2^+^ EVs from HC and AD in CSF and plasma showed the opposite trend, indicating the different role played by the blood-brain barrier (BBB) and the choroid plexus vascular barrier (PVB).

ROC analysis was then performed to evaluate diagnostic performance. In discriminating between HC and AD, the sensitivity and specificity for CCK^+^ EVs were 93% and 74% (AUC = 0.90, 95% CI 0.84∼0.96) (**Figure 5SJ**) and 65% and 83% (AUC = 0.78, 95% CI = 0.69∼0.86) for PMP2^+^ EVs (**Figure 5SL-O**), respectively. An integrative model combining these two biomarkers and the age factor discriminated AD from HC with an AUC of 0.95 (95% CI = 0.92∼0.99; sensitivity = 96%, specificity = 85%) (**Figure 5J**).

As for AD vs NAD, the sensitivity and specificity for CCK^+^ EVs were 76% and 72% (AUC = 0.78, 95% CI 0.69∼0.88) (**Figure 5SL**) and 64% and 47% (AUC = 0.53, 95% CI = 0.41∼0.65) for PMP2^+^ EVs (**Figure 5SM**). Again, an integrative model combining the two EVs markers and the age factor significantly improved the performance of the markers with an AUC of 0.84 (95% CI = 0.76∼0.92; sensitivity = 89%, specificity = 67%) (**Figure 5I**).

In summary, we chose the DEGs relative EVs as a promising biomarker to distinguish the AD from HC or NAD.

## Discussion

In this study, we have created a spatial atlas of gene expression in single-cell resolution for human hippocampus in both normal and diseased states. This atlas encompasses distinct molecular parameters, including integration of spatial mapping with alterations in the distribution of cells, genes, and functional pathways within hippocampal subregions and in relationship to Aβ plaques, reaching single-cell resolution. Furthermore, a select set of secretory markers identified as disrupted in AD has been validated within a sizable and well-defined cohort of both patients and healthy controls.

As the first molecular atlas depicting the hippocampal structure, it provides a detailed single-cell level guide to heterogeneous populations of cells, genes, and spatial structures, offering a molecular perspective for researchers. Using this atlas data, we have further constructed a whole-transcriptome atlas of the AD hippocampus. Comparative analysis between control and AD identified DEGs across the 14 subregions, reflecting the widespread, cross-regional alterations withing the AD hippocampus (**Figure 2A and Figure S2A**). In contrast, other DEGs were isolated to only one subregion, with CA4 standing out in the number of DEGs, thereby highlighting the vulnerability of this region in AD progression (**Figure 2B and Figure S2B**).

Furthermore, we also revealed changes in molecular pathway over various subregions of hippocampus. Notably, CA4 displayed more pronounced alterations in autophagy and metabolic pathways and differs from other CA in terms of protein folding (**Figure 2D**). Unsurprisingly, downregulation of many synaptic functions was observed, with synaptic pruning standing out in contrast as an upregulated system, indicating the possibility of abnormal synaptic pruning mediated by the complement system in microglia (**Figure 2D-H**). This phenomenon may lead to a significant loss of synapses, ultimately resulting in cognitive impairment. Another groundbreaking finding is the upregulation of genome genes within the electron transport chain and the genes coding proteins (MT.rcc), but not rRNA or tRNA (MT.rt), located in the mitochondrial nuclear DNA (**Figure 2I-J**). This discovery suggests that AD brains may employ a compensatory strategy at the metabolic level to address disruptions in hippocampal metabolism, enhancing mitochondrial function to meet the elevated energy demands of AD patients ^35^.

We delineated cellular architecture within hippocampus after joint analysis of snRNA-seq and Stereo-seq data, identifying cell types and their alterations within hippocampal subregions. The result enabled us to unravel spatial distribution of hippocampal cells and identify gene signatures corresponding to anatomical locations (**Figure 1**). Moreover, we have further uncovered pronounced changes of cell composition and gene expression in the FAS **(Figure 3)**, suggesting a relative higher inflammatory response in oligodendrocyte enriched area than neuron enriched area in the context of AD

In further characterizing changes specifically associated with AD, we considered the pathological hallmarks of accumulation of Aβ in plaques and NFTs with hyper-phosphorylated tau ^36–38^. In the atlas, we analyzed the distribution of the Aβ plaque, revealing that Aβ accumulation is prominent in the CA1, CA4 and SLRM of the hippocampus (**Figure 3A-B**). Statistical analysis of the cell density around the Aβ plaque suggests that the main cell types involved in response to accumulation of Aβ plaque are microglia and astrocytes, with microglia exhibiting more pronounced changes in proximity to the plaques (**Figure 3I-L**). Of note, in this study, we were not able to map out a gradient of gene expression in relationship to tau distribution due to unique expression profiles of NFTs in the tissue.

We next sought to demonstrate the utility of our atlas by exploring useful biomarkers for precise clinical diagnosis of AD. Among the altered pathways, we specifically extended our investigation to those related to vesicular transportation, reasoning that alterations that affect export to the plasma would be more plausible as peripherally accessible biomarkers, and would facilitate the application of our findings to clinical diagnosis and large-scale clinical screening. We found dramatic alterations in transport vesicles, endocytic vesicles, and the clathrin-coated vesicles in AD. Therefore, we compared the DEGs from our study to proteins previously identified in brain vesicles, as reported in the exobase database (http://www.exorbase.org/exoRBaseV2). Ninety-one brain-derived EV proteins were identified, and the cluster of the cell subtype distribution in the hippocampus were described in the secretion EVs map (**Figure 5D-E and Figure S5A-B**).

Having thus identified EV proteins with a high probability of AD-related alteration, we explored the possibility of using hippocampal specific markers to diagnose AD while differentiating it from various other types of dementias. Among several DEG proteins tested, we identified the EVs carrying CCK and PMP2 in plasma as robust diagnostic markers. Examination of their known functions shows their plausibility as relevant targets for alteration in AD. A recent study has reported that CCK, a satiety hormone, is highly expressed in several brain regions, especially the hippocampus, and is integral for maintaining or enhancing memory ^39^. PMP2 is known to be critical to the myelin of human central nervous system ^40,41^. Importantly, however, the exact roles and mechanisms of alterations of these proteins in AD are not known, demonstrating the power of the novel atlas as a system for investigation and discovery of additional aspects of this complex disease. Indeed, our data showed that the ratio of CCK^+^ and PMP2^+^ EVs not only accurately discriminated between AD patients and HC (AUC = 0.95) (**Figure 5H**), but also effectively distinguished AD patients from NAD (AUC = 0.84) (**Figure 5I**).

In conclusion, leveraging cutting-edge single-cell stereo-transcriptomics, we have constructed a comprehensive atlas of the human hippocampus, revealing novel pathological alterations unique to AD. Furthermore, we have identified promising biomarkers for AD diagnosis and differential diagnosis, laying the groundwork for future clinical applications.

## Methods

### Ethics statement

All participants signed an informed consent, an anatomical gift act for organ donation, and a repository consent to allow the data to be shared. All experiments related human brain tissues in this study were reviewed and approved by the Human Ethics Committee of the School of Medicine, Zhejiang University (approval no. 2022-012) and the Institutional Review Board on Ethics Committee of Beijing Genomics Institute (BGI, approval no. BGI-IRB 22080). Human plasma and CSF sample studies were approved by the clinical research ethics committees of the First Affiliated Hospital, College of Medicine, Zhejiang University (approval no. 2022-043).

### Brain tissue collection

Human brain tissue from the middle of hippocampus were obtained from Brain specimens were obtained from the National Human Brain Bank for Health and Disease of China in Zhejiang University (CNBB). The samples were assigned to groups based on both NFTs and Aβ plaque staging, in addition to clinical diagnoses. Samples were also selected based on several covariates, including age, sex, race, postmortem interval (PMI), RIN, and disease comorbidity. To assess RNA quality, two tissue samples (approximately 50 mg each) were collected from the tissue slab corresponding to both the frontal pole and temporal pole of each donor’s brain. The dissected tissues were preserved in 1.5 mL microcentrifuge tubes, either on dry ice or at the −80 ℃, until the RNA isolation. The isolation of RNA was performed using the Qiagen RNeasy Mini Kit (Qiagen, 74104), following the manufacturer’s instructions. The determination of RIN values for each sample was carried out using the Agilent RNA 6000 Nano chip kit (Agilent, 5067-1511) and an Agilent Bioanalyzer 2100 instrument, following the manufacturer’s provided protocol. Only samples with RIN >7.0 were applied in this research. Sample information is available in **Table 1**.

### Cohort of participants and sample collection for exosome application

The procedures, including standard protocol approvals, registrations, and patient consents, garnered approval from the institutional review boards at the first affiliated hospital, Zhejiang University school of medicine, Hangzhou, China.

Plasma samples were comprised 45 subjects diagnosed with AD, 43 subjects with Non-AD dementia (NAD), and 66 healthy controls (HCs), who were enrolled at the first affiliated hospital, Zhejiang University school of medicine (**Table 2**). Using the molecular signature of Positron Emission Tomography (PET) to select individuals with ’true’ AD and HCs, 45 with AD and 66 HCs met the previously identified diagnostic criteria ^42–44^. All participants underwent a comprehensive clinical evaluation, and the criteria for inclusion, exclusion, and the procedures for sample collection have been previously described ^45–49^.

**Table 2.**
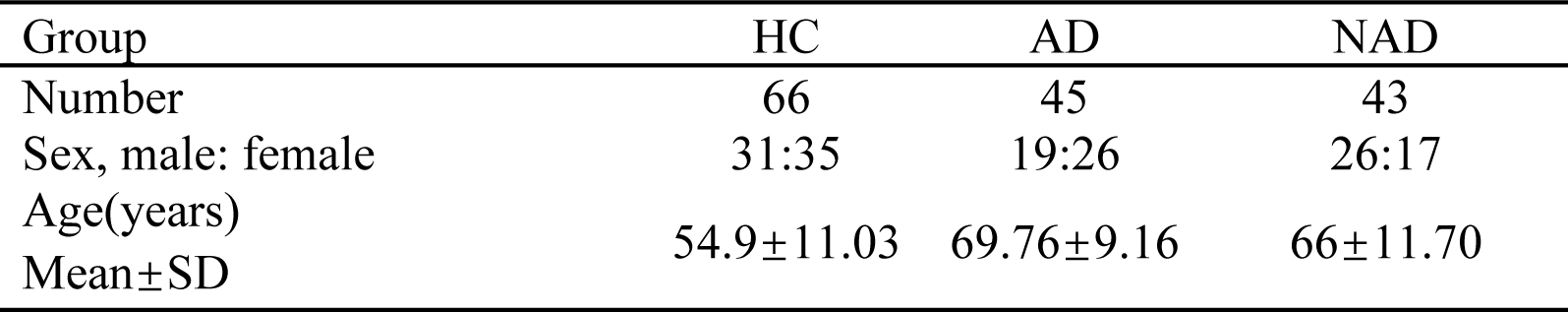
Summary of the demographics and clinical data of participants.

Reference plasma samples pooled from 30 healthy controls, were obtained from the first affiliated hospital, Zhejiang University school of medicine, as mentioned previously ^45,46^ ^50^.

### Stereo-seq chip preparation

Capture chips were created following the Stereo-seq protocol ^51^. Briefly, to establish the DNB array for in situ RNA capture, we initially synthesized oligonucleotides containing random 25-nucleotide coordinate identity (CID). These oligonucleotides were circularized using T4 DNA ligase and splint oligonucleotides. Subsequently, DNBs were produced through rolling circle amplification and placed onto the designed chips (65 mm × 65 mm). To identify unique DNB-CID sequences at each spatial position, single-end sequencing was conducted using a DNBSEQ-Tx sequencer (MGI Research, Shenzhen, China) employing the SE25 sequencing strategy. After sequencing, oligonucleotides containing poly-T and a 10-bp Molecular Identity (MID) were hybridized and ligated to the DNB located on the chip. This capture probes generated with a 25 bp CID barcode, a 10 bp pair MID, and a 22 bp poly-T, prepared for in situ capture. CID sequences, along with their respective coordinates for each DNB, were identified using a base calling method as per the manufacturer’s instructions for the DNBSEQ sequencer. Following sequencing, the capture chip was divided into smaller chips (10 mm × 10 mm), eliminating any duplicated CIDs in non-adjacent positions.

### Stereo-seq tissue processing and imaging

The Stereo-seq experiments were conducted as previously described ^17,52^. The Stereo-seq capture chip was washed with NF-H_2_O containing 0.05 U/µL RNase inhibitor (NEB, M0314L) and the chips were dried at room temperature. The cut hippocampus tissue sections were adhered onto the Stereo-seq capture chip surface and incubated at 37°C for 3 minutes. Subsequently, the sections were fixed with methanol and stored at −20°C for 40 minutes before assembling Stereo-seq library. After fixation, chips with sections were stained with a nucleic acid dye (Thermo Fisher, Q10212), and imaging was taken by using a Ti-7 Nikon Eclipse microscope prior to *in situ* capture at the FITC channel (objective 10×).

### Stereo-seq *in situ* reverse transcription

After washing the sections with 0.1×SSC wash buffer (Thermo, AM9770) containing 0.05 U/ml RNase inhibitor (NEB, M0314L), the chip coated with tissue sections was permeabilized with 0.1% pepsin (Sigma, P7000) in 0.01 M HCl buffer, incubated at 37°C for 12 minutes and then washed with 0.1×SSC wash buffer containing 0.05 U/mL RNase inhibitor (NEB, M0314L). The RNA released from the permeabilized tissue and captured by the DNB was reverse transcribed overnight at 42°C. This process used SuperScript II (Invitrogen, 18064-014) containing10 U/uL reverse transcriptase, 2.5 mM Stereo-seq-TSO (5-CTGCTGACGTACTGAGAGGC/rG//rG//iXNA_G/-3), 1 x First-Strand buffer, 7.5 mM MgCl_2_, 5 mM DTT, 2 U/ml RNase inhibitor 1 mM dNTPs and 1 M betaine solution PCR reagent. Following the reverse transcription step, the tissue was digested with Tissue Removal buffer (25 mM EDTA,10 mM Tris-HCl, 0.5% SDS, 100 mM NaCl) at 37°C for 30 minutes. The cDNA-contained chips were then subjected to Exonuclease I (NEB, M0293L) treatment for 1 hour at 37°C and were finally washed twice with 0.1×SSC wash buffer.

### Amplification

The obtained first-strand cDNAs were subjected to amplification using KAPA HiFi Hotstart Ready Mix (Roche, KK2602) along with a cDNA-PCR primer(5-CTGCTGACGTACTGAGAGGC-3) at a concentration of 0.8 μM. The PCR reactions were carried out as described below: an initial incubation at 95°C for 5 minutes, followed by 15 cycles of denaturation at 98°C for 20 seconds, annealing at 58°C for 20 seconds, extension at 72°C for 3 minutes, and a final step at 72°C for 5 minutes.

### Library construction and sequencing

The resultant PCR products were purified by using VAHTS DNA Clean Beads (Vazyme, N411-03, 0.6 ×), and then quantified by Qubit^TM^ dsDNA Assay Kit (Thermo, Q32854). After this process, a total of 20 ng of cDNA was then fragmented by employing in-house Tn5 transposase at a temperature of 55°C for 10 minutes. After the fragmentation was completed, the reactions were halted through the addition of 0.02% SDS and gently mixing it at room temperature for 5 minutes. According to this, the fragmented products entered the amplification phase. The PCR products were amplified as follows: a 25 μL PCR product was mixed with 1× KAPA HiFi Hotstart enzyme. The mixture included 0.3 μM of Stereo-seq-Library-F primer (5’-CTGCTGACGTACTGAGAGG*C*A-3’) and 0.3 µM of Stereo-seq-Library-R primer (5-GAGACGTTCTCGACTCAGCAGA-3’). The total reaction volume was 100 μL, including the addition of NF-H_2_O. The amplification process consisted of the following steps: 1 cycle at 95°C for 5 minutes, 13 cycles at 98°C for 20 seconds, 58°C for 20 seconds, and 72°C for 30 seconds, and a final cycle at 72°C for 5 minutes. Subsequently, the amplified products were purified using AMPure XP Beads, with purification volumes of 0.6 × and 0.15 × the reaction volume. These purified products were then used to generate DNBs (DNA nanoballs). Finally, the prepared DNBs underwent sequencing on the MGI DNBSEQ-Tx sequencer at the CNGB.

### Immunohistochemical (IHC) and immunofluorescent (IF) staining

The hippocampus sections were cut at 10 µm and then fixed by immersion in methanol for 30 minutes at −20℃. Subsequently, they were washed with TBST (Tris Buffered Saline with 0.25% Tween) twice for 3 minutes each. The slides were blocked using a 5% NGS blocking solution (Jackson Immunoresearch) at 4℃ for 1 hour. Following this, the slides were incubated with the antibodies at 4℃ overnight. The following primary antibodies were used at the appropriated dilutions: β-Amyloid (1:200, 6F/3D Mouse, DAKO, M0872), PHF-TAU (1:1000, AT8, Mouse, Thermo Fisher, MN1020), Iba-l (1:500, Rabbit, Wako, 019-19741), GFAP (1:500, Chicken, Abcam, 134436). After the primary antibody incubation, the sections were washed 3 times for 5 minutes each with TBST, and then stained with a horseradish peroxidase-conjugated secondary antibody (Proteintech) or Alexa Fluor-conjugated secondary antibody (Invitrogen). Subsequently, the sections were washed 3 times for 5 minutes each with TBST, and the antibody complex was visualized by HRP-mediated oxidation of 3,3’-diaminobenzidine (DAB) for the IHC, and counterstaining was performed with hematoxylin after the DAB reaction. Finally, cover slipping was carried out using mounting medium (Solabrio) or the antifade mounting medium with DAPI (H-1400, Vector Laboratories).

To analyze the different slides obtained from the hippocampus tissue samples processed for IHC, image acquisition of the tissue samples was performed using a digital scanner (Kifbio). For the IF, the visualized using a fluorescence microscope (Stellaris8 Falcon, Leica).

### Hippocampus tissue collection for snRNA-seq

To remove a specific region of interest from frozen brain slabs, which were 500 μm thick and intended for downstream nuclear sequencing applications, the tissue slabs were taken out of storage at –80°C, briefly transferred to a –20°C freezer to prevent tissue shattering during dissection, and then carefully handled on a custom freezing microtome maintained at –20°C during dissection. Dissections were performed using razor blades or scalpels cooled by dry ice to prevent tissue warming. The dissected tissue samples were transferred to cryogenic storage vials, sealed, and stored at −80°C until they were needed.

### Single-nucleus suspension preparation

The procedure for single-nucleus isolation was executed following the method described previously ^53^. Briefly, the tissues were thawed, minced, and transferred into a 1-mL Dounce homogenizer (TIANDZ), along with 1 mL of homogenization buffer A. The buffer consisted of 250 mM sucrose (Ambion), 10 mg/mL BSA (Ambion), 5 mM MgCl_2_ (Ambion), 0.12 U/μL RNasin Plus (Promega, N2115), 0.12 U/μL RNase inhibitor (Promega, N2115), and 1 × Complete Protease Inhibitor Cocktail (Roche, 11697498001). The frozen tissues were kept in an icebox and homogenized using the loose pestle (pestle 1) for 25-50 strokes. Subsequently, the mixture was passed through a 100-µm cell strainer into a 1.5-mL tube (Eppendorf). After that, 750 μL of buffer A, which included 1% Igepal (Sigma, CA630), was added to a clean 1-mL Dounce homogenizer. The tissue was further homogenized using the tight pestle (pestle 2) for 25 strokes. Following filtration through a 40-µm strainer into a 1.5-mL tube, the mixture was centrifuged at 500 *g* for 5 min at 4 °C to form a pellet of nuclei. The pellet was subsequently resuspended in 1 mL of buffer B, which consisted of 320 mM sucrose, 10 mg ml/L BSA, 3 mM CaCl_2_, 2 mM magnesium acetate, 0.1 mM EDTA, 10 mM Tris-HCl, 1 mM DTT, 1× Complete Protease Inhibitor Cocktail, and 0.12 U μl/L RNase inhibitor. The resuspended nuclei were then centrifuged again at 500 *g* for 5 min at 4 °C to form another pellet of nuclei. Finally, these nuclei were resuspended in a cell resuspension buffer at a concentration of 1,000 nuclei per μL for library preparation.

### snRNA-seq library construction and sequencing

The DNBelab C Series Single-Cell Library Prep Set (MGI, 1000021082) was employed as previously described ^54^. In summary, single-nucleus suspensions were utilized to create barcoded libraries through droplet generation, emulsion breakage, bead collection, reverse transcription, and cDNA amplification. Indexed libraries were prepared following the manufacturer’s protocol, and their concentrations were determined with a Qubit ssDNA Assay Kit (Thermo Fisher Scientific, Q10212). The libraries were sequenced on either a DNBSEQ-Tx sequencer at the China National GeneBank (CNGB, Shenzhen, China), with a sequencing strategy of 41-bp read length for read 1 and 100-bp read length for read 2.

### snRNA-seq data processing

Raw sequencing reads from DNBSEQ-Tx were aligned to the GRCh38.p12 genome. The snRNA-seq profiles nuclear precursor mRNA (pre-mRNA), which includes transcripts that have not completed splicing to remove introns. Inclusion of intron readings from pre-mRNA in the final gene expression count enables capture of all the information in the pre-mRNA. Thus, each gene’s transcript in snRNA-seq was counted by including exon and intron reads together. We used SoupX (v1.6.2;https://github.com/constantAmateur/SoupX) to reduce ambient RNA noise. The rho value was automatically parameterized using the “autoEstCont”function in tissues.

### Quality control and cell clustering of snRNA-seq dataset

The quality control and clustering analysis of the snRNA-seq dataset was performed using Seurat (version 4.2.2) and the R program. In detail, we first identified and removed the doublets using DoubletFinder (version 2.0.3). Low-quality cells with less than 500 detected genes or more than 98% of cells in gene number in each library, as well as cells with the mitochondrial gene ratio of more than 10% in data preprocessing, were filtered out, and all query genes were guaranteed to be expressed in at least three cell prior to further use. The top 3000 highly variable genes were selected according to the “FindVariableFeatures” function. For downstream clustering and visualization, principal component analysis (PCA)-based dimension reduction was initially performed, and the first 30 principal components (PCs) were extracted for subsequent Louvain clustering to define the cell clusters (the resolution was set to 0.5). The clustering results were finally characterized in a two-dimensional space using the UMAP, and the cell clusters were annotated using known biomarkers that were more highly expressed in a particular cluster (via the FindAllMarkers function with parameters: *p*.adjust < 0.05, avg_log_2_FC > 0.25, min.pct = 0.1).

### Stereo-seq data bin100 processing

The spatial transcriptomic gene expression profile matrix was divided into non-overlapping bins covering an area of 100×100 DNB (Bin100). The UMIs were aggregated per gene within each bin. The resulting Bin100 expression profile matrix containing coordinates were further used to generate Seurat object in Seurat package ^55^ in R (version 4.2.2). The NormalizeData function with default parameters was used to erase the depth differences of all bins.

### Subregion parcellation of hippocampus

To achieve an accurate brain region parcellation, we integrated adjacent sections of H&E staining images and spatial transcriptomic data. Initially, pathologists divided the anatomical structures based on cell morphology, density, and staining intensity in the HE staining images, providing a delineation of brain regions. Then, the transcriptomic profiles of divided bins (Bin100) on every chip were used as input to generate a standardized SingleCellExperiment object for subsequent BayesSpace ^56^ clustering. Principal component analysis (PCA) was performed on the top 3000 highly variable genes, and the top 50 principal components were retained. Clustering was performed with a maximum cluster count over 10,000 iterations. The two-dimensional brain region contours obtained from H&E staining were extracted using an image processing R package (imager: http://dahtah.github.io/imager/). After registering the contours with the spatial clustering results, refinement of the contour lines was performed based on the BayesSpace clustering of spatial bin100 data. Then, the parcellation of hippocampal subregions were generated with integration of morphological and Stereo-seq data.

### Multiple-sample integration and batch correction

To perform cross-sample analysis, we integrated data from six individual samples. Prior to integration, data of each sample underwent separate data preprocessing. This includes quality control, filtering out low-quality cells, and normalizing the data. After merging the data using the merge function, dimensionality reduction through PCA and NormalizeData function were consistently applied to the integrated dataset. Subsequently, batch correction was performed using the Harmony ^57^ package (https://github.com/immunogenomics/harmony).

### Dimension reduction and clustering

All the dimensionality reduction analyses used a combination of PCA and uniform manifold approximation and projection (UMAP). PCA, a linear dimensionality reduction method, was employed to extract primary features by reducing redundancy in the data. UMAP, a nonlinear dimensionality reduction technique, was used to capture the complex structures and subpopulations within the data. For the batch-corrected integrated data, UMAP clustering is typically performed using 1-30 harmony reduction. Unsupervised clustering was performed using the FindNeighbors and FindClusters functions from the Seurat package.

### Identification of hippocampus region-specific markers in normal individuals

To identify representative markers for each brain region, we first converted the region delineation obtained earlier into group information for each bin and added it to the metadata. In this section, we utilized data from all six chips belonging to the control group as input. The FindAllMarkers function was employed to individually identify markers for each brain region in every chip. We used the Wilcoxon rank-sum test (default) based on the normalized values, and the genes with *p*-value < 0.05 and log_2_FC threshold = 0.25/0.2 were selected as region highly expressed genes. Highly expressed genes that were specifically expressed in a single brain region and repeated in the corresponding regions of all six chips were defined as region-specific markers.

### Gene set analysis for Stereo-seq bin100

The pre-defined gene sets were collected from Gene Set Enrichment Analysis (GSEA) and KEGG (Kyoto Encyclopedia of Genes and Genomes) databases. These gene sets represented known biological pathways or functional groups and have been manually curated and classified to remove pathways with similar meanings and redundant gene compositions. We utilized the AUCell package to score these gene sets in both the AD and control groups. Initially, the standardized expression matrix was transformed into a rank matrix. Subsequently, we calculated the area under the recovery curve, setting the activity threshold for each gene set at 5%.

### Spatial expression pattern analysis

To quantify and analyze the spatial distribution of transcriptomic features, the CA1 and SLRM regions were separately extracted and subjected to a more detailed regional division. For SLRM, we defined the orientation from CA1 to DG as the CA1-DG axis and mapped this axis to a continuous variable ranging from 1 to 5 layer. As SLRM comprised five layers along the CA1-DG axis, we directly computed the average expression of each gene in these five layers and fitted a linear model using the continuous variable on the CA1-DG axis. The positive or negative slope of the linear model indicated whether gene expression increases or decreases along the CA1-DG axis.

For CA1, we defined the orientation from CA2 to CA1 as the CA2-CA1 axis. Firstly, the entire CA1 region was divided into 3-6 blocks, with each block assigned an angle. For bin100 unit *a* (*x,y*) with assigned angle *b:*

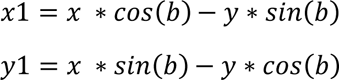

The coordinates (*x1, y1*) represented the transformed position of bin100 unit *a*. After applying coordinate transformation to all spots within the CA1 region, along the CA2-CA1 axis, CA1 was divided into multiple columns at intervals of 500um and all the columns were mapped to continuous variables ranging from 0 to 1. Then we computed the average expression of each gene in these columns and fitted linear model using the continuous variable on the CA2-CA1 axis. The positive or negative slope of the linear model indicated whether gene expression increases or decreases along the CA2-CA1 axis.

### Image-based single-cell segmentation of stereo-seq data with StereoCell

AI-assisted cell segmentation was performed to our Stereo-seq data as described previously ^58^. Simply, we summed the total UMI counts within each DNB point corresponding to specific spatial coordinates to form a spatial density matrix, which was then transformed into an image representation where each pixel in the image represented a DNB point, and its grayscale intensity reflected the total UMI count. We projected the nucleic acid staining image to the Stereo-chip data for the same tissue section by manually registration. Cell segmentation was performed using the proposed model trained and optimized based on ESPANet (https://arxiv.org/abs/2105.14447), aiming to improve multi-scale representation capabilities at a finer granularity and develop long-range channel dependencies. This was followed by post-processing using a watershed algorithm, ultimately generating black-and-white mask results. For each segmented cell, UMIs from all DNBs within the corresponding segment were aggregated per gene and then summed to create a cell-by-gene matrix for downstream analysis.

### Cell type annotation of segmented cells by Spatial-ID

Spatial-ID algorithm was performed to annotate segmented cells in Stereo-seq as described previously ^27^, with some modifications. Initially, a four-layer deep neural network (DNN) trained on snRNA-seq data computed initial probabilities for cell types in Stereo-seq. Subsequently, spatial relationships among cells were encoded into an adjacency matrix with normalized Euclidean distances. A graph convolution network (GCN) incorporated the initial probabilities, adjacency matrix, and gene expression profiles, employing two autoencoders for gene and spatial representations, and a classifier block. The GCN underwent xxxepochs, emphasizing self-supervised learning for feature extraction and Cross Entropy loss for refining cell type probabilities. Cell types were determined by the highest GCN-generated probability. Implemented using PyTorch(v.1.13.1) and PyTorch Geometric(v.2.1.0) in Python (v.3.8.10), spatial-ID mitigated batch effects across Stereo-seq sections during cell type annotation.

### Quality control of single-cell Stereo-seq data

Processing of the MID count matrix obtained from Stereo-seq data was implemented using the R package Seurat (v4.0.0). To remove low-quality cells, we filtered out cells with gene numbers no more than 100 for all cell types. Furthermore, we discarded low-quality cells with a high percentage of mitochondrial genes (> 20%). After filtering, the remaining 701,092 single cells with 293 genes were included in the downstream analyses.

### Correlation analysis of cell types between stereo-seq and snRNA-seq data

The snRNA-seq and Stereo-seq data were normalized using the R package Seurat, with reformatting of the SCT assay slot. Within this slot, the top variable genes were included and encompassed for subsequent analysis. Overlapping top variable genes, present in both the snRNA-seq and Stereo-seq datasets, were utilized to compute Pearson correlations. The primary objective of this methodology was to ascertain the correspondence of cell types between the snRNA-seq and Stereo-seq datasets.

### Neighborhood complexity analysis

The neighborhood complexity analysis was performed using squidpy (1.3.0) ^59^. The connectivity matrix was computed for each section by itself and then compiled by calculating mean scores. Cluster proximity of cell type is quantified with a permutation-based test (1000 permutations), by comparing the actual cell type labels with a random configuration of labels and maintaining the positional information. From each pair (actual label vs label of each permutation), the means and standard deviations are estimated, and a Z-score is calculated.

### Identification of DAM and DAA within microglia and astrocytes

We integrated microglia and astrocytes in our data using the Harmony package. Subsequently, principal component analysis (PCA) was performed, selecting the top 30 principal components for downstream analysis. Following that, the integrated dataset was re-clustered using the “FindNeighbors” and “FindClusters” functions, with a clustering resolution set at 0.6. Finally, by employing the ’AddModuleScore’ function from Seurat (version 4.2.2), the subcluster with the highest scores was annotated as either DAM or DAA, utilizing the specific gene sets allocated for DAM or DAA.

### Registration and analysis of Aβ staining images

Imager and imagerExtra packages in R were employed for the registration of Aβ staining images with spatial transcriptomic images. The essence of registration is to align the features of the two images. If the features to be registered in spatial transcriptomic image was obscure, we added the feature markers manually to feature sites of two aligned images. After alignment, we extracted the coordinates of stained Aβ plaques from the image and registered to the coordinates of the spatial transcriptomic image. Each plaque coordinate consisted of hundreds to thousands of pixels, which can be used to reconstruct the outline of the plaque. Each plaque represented a point cloud and the alphashape algorithm (XXX) was used to identify boundary points of the plaque. Subsequently, these boundary points were sorted using polar coordinates and then generated a polygon objects for gaining the boundary of the plaque.

### Identification of Aβ plaque-related genes

To identify Aβ plaque-related genes, we first determined the coordinate of each plaque center which can be defined by calculating mean value of x and y axis, respectively, from a point cloud of the plaque. Then, taking plaque center as center point, we constructed an extended square with 150 μm in side around the center point. This kind of squares with Aβ plaque was designated as AD-Aβ squares. Simultaneously, in the same slice, we randomly delineated a series of squares of same dimension size with AD-Aβ square in subregion without accumulation of Aβ plaques. This kind of squares in AD slice was designated as AD-non-Aβ squares. Finally, in control slice, we randomly delineated a series of squares of same dimension size in area corresponding to Aβ enriched region in AD slice. This kind of squares in control was designated as control squares. Based on above definition, we categorized squares into 3 groups including, AD-Aβ group, AD-non-Aβ group and control group. DEG analysis was first performed between the AD-Aβ group and the control group. Subsequently, DEG analysis was conducted between the AD-Aβ group and the AD-non-Aβ group. The intersection of the two sets of DEGs were conducted and these genes after intersection were defined as Aβ plaque-related genes.

### Analysis of cell type distribution density around the Aβ plaque

Figure 4L calculated the density distribution of cell types at different distance intervals (0-20, 20-40, 40-60, 60-80 and 80-100 μm) to the Aβ plaque. The density distribution was obtained by using the sp (SpatialPolygons: version 1.5-1) package in Rstudio. Firstly, the boundary points of each plaque were transformed into the raw spatial polygon objects. Secondly, we used the “gContains” function to identify cells located within these plaque polygons objects, and simultaneously utilized the “gArea” function to obtain the area of these plaques. For example, in plaque A, we calculated the density of various cell types in plaque A. As a result, we got the average density of various cell types under plaques. Subsequently, we equidistantly extended 20 μm from the boundary of each plaque to generate new polygons objects. Subtracting the area of new polygons object from the raw object’s area, we obtained the area of the first ring. Cells within this ring were also identified using the “gContains” function. In this way, we could obtain the density of various cell types within the 0-20 μm range around the plaque. Repeating above steps, we obtained the density of different cell types at various distance around the plaque.

### AD-associated DEG analysis in snRNA-seq and Stereo-seq data

We performed AD-associated DEG analyses using the “FindMarkers function” in Seurat package with Stereo-seq and snRNA-seq data, and without specific mentions, the parameters were set as following: *p*-value < 0.05, |log_2_FC| > 0.25 and min.pct = 0.1. In this study, only genes expressed more than 15% of cells or bin100s were regarded as DEG. Region-specific DEGs indicate that a gene was defined as differentially expressed only in a particular region. Shared DEGs imply that the gene exhibits differential expression simultaneously in multiple brain regions.

### Ontology annotation

Website tool DAVID (https://david.ncifcrf.gov/tools.jsp) and ClusterProfiler package in R were used for the Gene Ontology (GO) term enrichment. Simplify function with ‘‘cutoff = 0.6’’ parameter was used to remove the redundant GO terms. GO terms with *p*-value < 0.05 were defined as significantly enriched.

### Cryo-Electron Microscopy (Cryo-EM)

EVs (5 μL) were deposited on an EM grid coated with a perforated carbon film and incubated for 30 minutes; the liquid was blotted from the back side of the grid and the grid was quickly plunged into liquid ethane using a Leica EMCPC cryo-chamber. EM grids were stored under liquid nitrogen prior to EM observation. Cryo-EM was performed with a Titan Krios electron microscope (ThermoFisher, USA).

### Plasma sample processing and detection by CytoFLEX

Plasma samples were rapidly thawed at 37°C and centrifuged at 3000 *g* for 30 minutes at 4°C, followed by centrifugation at 10,000 *g* for 30 minutes at 4°C. 10 μL of clarified plasma samples were diluted with 70 μL of phosphate-buffered saline (PBS; 0.22μm-filtered) and 20 μL of 40% PEG8000). The mixture was thoroughly vortexed and allowed to incubate at room temperature for 30 minutes. Subsequently, it was centrifuged at 12,000 *g* for 20 minutes at 4°C. The pellets were resuspended in 100 μL of filtered PBS and stored at −80°C until use.

For EVs analysis, 10 μL of EVs were blocked with an equal volume of 2% BSA (Bovine Serum Albumin), followed by labeling with 0.03 μg of fluorescently tagged antibodies. The samples were then incubated at 4°C in the dark overnight. On the following day, 62 μL of 0.22 μm-filtered PBS and 5 μL of a lipid probe (10 μM) were added to the samples, resulting in a final reaction volume of 100 μL with a lipid probe concentration of 500 nM. The samples were incubated at 4°C in the dark for 1 hour. Subsequently, 100 μL of 4% PFA (Paraformaldehyde) was added for fixation, and the samples were fixed for 20 minutes. After appropriate dilution, the samples were analyzed using a CytoFLEX flow cytometer (VSSC mode), a total of 50,000 particles were collected under the lipid probe gate.

### Nanoparticle tracking analysis (NTA)

NTA was executed using a NanoSight NS300, following the manufacturer’s guidelines. In breif, 10 μL of PEG8000-treated reference plasma underwent a 1:100 dilution with 0.22 μm-filtered PBS (pH 7.4), ultimately resulting in a final volume of 1 mL for direct scatter measurements. For each assessment, fraction, three repeated videos (60 s each) were captured for analysis. Subsequently, these videos were analyzed using NanoSight Software NTA 3.2 software (Nanosight, Amesbury, UK).

### Statistical analyses

For EV analysis, all analyses were performed in SPSS 25.0 (IBM) or Prism 8.0 (GraphPad Software). ROC curves for analytes were generated to evaluate their sensitivities and specificities in distinguishing AD from HCs or NAD. Logistic regression was used to generate an integrative model that included the two plasma biomarkers. P < 0.05 was regarded as significant

## Data Availability

All data produced in the present study are available upon reasonable request to the authors

## Acknowledgments

Study data were generated from postmortem brain tissue obtained from the CBB (National Human Brain Bank for Health and Disease) University of Zhejiang, which is supported by the Ministry of Science and Technology Project Subsidy Project for the National Health and Disease Brain Tissue Resource Bank, Zhejiang Provincial Science and Technology Department Key Project, Construction of the National Health and Disease Brain Tissue Resource Bank grant 2020ZY1020. This work was supported by National Natural Science Foundation of China under Grant 82020108012, Leading Innovation and Entrepreneurship Team of Zhejiang Province under Grant 2020R01001, and Innovative Institute of Basic Medical Science of Zhejiang University and the National Science and Technology Innovation 2030 Major Program (2021ZD0204401). We thank BGI for hosting various datasets from this study.

## Author contributions

P.W, LF.W, and J.Z conceptualized and designed the project; T.L, MN.C, YJ.J, performed and analyzed the sequencing data; L.H, J.W and X.X supervised the work; P.W, Z.X and J.Z designed the experiments; ZQ.F, PR.J, JL.W, B.S, F.S, BY.L, GP.P, XH.S, C.P, Y.S, AM.B and KQ.Z collected tissue samples; Z.X, Z.G, JY.Y, C.T, B.X, Y.Y, SP.W, and T.L performed the experiments. QY.T, YZ.H, YY.L, Y.G, ZH.W,BF.J, HW.Z, YR.W, JJ.L, HY.W,YC.T, SP.L and JH.Y performed data analysis. P.W, Z.X, Z.G and LF.W prepared the figures. XM.L, LQ.L, H.L and SM.D provided relevant advice and reviewed the manuscript. P.W, LF.W, QY.T, Z.G, T.L, Z.X. and J.Z wrote the manuscript with input from all authors. All other authors contributed to the work. All authors read and approved the manuscript for submission.

## Additional information

### Supplementary information

Correspondence and requests for materials should be addressed to Jing Zhang, Lei Han, Shumin Duan and Longqi Liu.

